# Triage Administration of Ondansetron for Gastroenteritis in children; a randomized controlled trial

**DOI:** 10.64898/2026.04.13.26350796

**Authors:** Olivia Weill, Nathalie Lucas, Benoit Bailey, Christopher Marquis, Jocelyn Gravel

## Abstract

**Objectives:** Acute gastroenteritis is a leading cause of pediatric emergency department (ED) visits. While ondansetron reduces vomiting, intravenous rehydration, and hospital admissions, its efficacy when initiated at triage remains unclear. We aimed to evaluate whether triage nurse-initiated administration of ondansetron in children with suspected gastroenteritis reduces the proportion of patients requiring observation following initial physician assessment.

**Methods:** We conducted a randomized, double-blind, placebo-controlled trial in a tertiary pediatric ED in Canada. Children aged 6 months to 17 years presenting with morae than 3 episodes of vomiting in the preceding 24 hours (including 1 within 2 hours of arrival), were eligible. At triage, we randomized participants to receive liquid ondansetron or a color- and taste-matched placebo. The primary outcome was the proportion of patients requiring observation after the first physician evaluation. Secondary outcomes included post-intervention vomiting, ED length of stay, patient comfort, and 48-hour return visits. The trial was registered at ClinicalTrials.gov (NCT03052361).

**Results:** Recruitment was stopped prematurely due to the COVID-19 pandemic. Ninety-one participants were randomized to ondansetron (n= 44) or placebo (n= 47). Overall, 40 patients (45%) were discharged immediately after the initial physician assessment, with no difference between the ondansetron and placebo groups (44% vs. 45%; absolute difference -1%, 95% CI: -20% to 19%). No significant differences were observed in all secondary outcomes.

**Conclusion:** In this trial, triage nurse-initiated ondansetron administration did not reduce the need for ED observation in children with presumed gastroenteritis. While being underpowered, this study could inform researchers planning larger clinical trials.

## Introduction

Acute gastroenteritis (AGE) is one the most common cause for pediatric visits and hospitalizations worldwide with children under five years of age experiencing one to two episodes annually in industrialized nations(1-3). In the United States, AGE accounts for 20% of outpatient visits among younger children, and more than 200,000 hospitalizations per year(4). Although most cases are self-limiting and viral in origin, persistent vomiting may results in dehydration and failure of oral rehydration, thereby requiring intravenous (IV) fluids or hospital admission(5).

Multiple clinical trials have established that oral ondansetron effectively reduces the need for IV fluids, length of stay and hospital admissions in children with AGE(6-10). It was also demonstrated that there is no risk of arrhythmia following short term administration of oral ondansetron unless the patient had a known risk of ventricular arrhythmia(6, 11).

The administration of ondansetron at triage, rather than after physician assessment, could theoretically expedite recovery, improve patient comfort, and reduce the need for prolonged ED observation. While some centers have implemented triage-based protocols for antiemetics, high-quality prospective data are lacking. A cross sectional survey conducted in Ontario, Canada in 2010 reported that 16% of 133 EDs had a standardized directive for the use of an antiemetic at triage(12). Another study reported the implementation of a triage-based protocol for oral rehydration and ondansetron triage(13). This quality improvement initiative improved the use of oral rehydration and ondansetron administration and decreased the need for intravenous fluid. To our knowledge, there is no prospective trial evaluating the effectiveness of ondansetron provided at triage. The primary objective of this study was to determine whether triage-initiated administration of ondansetron in children with suspected AGE reduces the proportion of patients requiring observation after the initial physician assessment. Secondary objectives were to evaluate its effect on patient comfort, emergency department (ED) length of stay, and return visits to a physician within 48 hours.

## Materials and Methods

We conducted a randomized, double-blind, placebo-controlled trial between October 2018 and March 2020 in a tertiary pediatric ED with an annual census >80,000 visits.

Children aged 6 months to 17 years weighing ≥8 kg were eligible if they had symptoms suggestive of AGE, including ≥4 episodes of non-bilious, non-bloody vomiting in the preceding 24 hours and ≥1 episode within 2 hours of arrival. At triage, the evaluating nurse was required to identify AGE as the most likely diagnosis. Exclusion criteria included severe dehydration according to the triage nurse, bloody diarrhea, history of abdominal surgery, known allergy to ondansetron or any of its components, long QT syndrome or significant cardiac disease, previous enrollment in the study, or inability to obtain parental informed consent (e.g., language barrier or absence of a legal guardian).

The study protocol was approved by the institutional review board and authorized by Health Canada (HC233368). The trial was registered at ClinicalTrials.gov (NCT03052361). Written informed consent was obtained from a parent or legal guardian, and assent was obtained from the child, when appropriate, before any study procedures were performed.

Participants were randomized to ondansetron or placebo allocated in the ED triage. Posology of ondansetron was adapted to weight (2 mg for 8–15 kg; 4 mg for 15–30 kg; 8 mg for >30 kg). Patients allocated to placebo arm received an identical looking and tasting placebo. Composition of placebo was sucrose 4,4 g/5 mL. In both arms: after 30 minutes of the administration of the intervention, oral rehydration was initiated according to a standardized treatment protocol currently used in the ED: 15 mL of preferred oral rehydration solution every 15 minutes continued until the initial physician assessment. Because it was standard ED practice to administer ondansetron to children actively vomiting at the time of physician assessment, withholding antiemetic therapy for those randomized to placebo was considered unethical. Accordingly, children for whom the treating physician deemed antiemetic therapy necessary after examination were offered a rescue dose, consisting of the alternate study medication (crossover), using the same dosing and administration protocol.

The primary outcome was binary (success vs. failure). Success was defined as discharge immediately after the initial physician assessment, without the need for ED observation. This outcome was selected by consensus among pediatric emergency physicians at the study site, who considered it clinically meaningful and less susceptible to the effects of ED crowding than total length of stay. Secondary outcomes included ED length of stay (from registration to discharge), number of vomiting episodes in the ED after the intervention, number of vomiting episodes within 24 and 48 hours, and the proportion of patients returning to the ED or to another physician within 48 hours. Patient comfort was evaluated by parents according to the Baxter Animated Retching Face (BARF) scale(14) for patients older than 7 years old.

As part of the safety analysis, we assessed whether the triage nurse’s initial suspicion of AGE was confirmed at the end of the ED visit and compared the proportion of alternative diagnoses between groups. We also compared the proportion of children who left the ED without being seen by a physician and the proportion who required rescue antiemetic medication for persistent nausea or vomiting.

Participants were randomized in a 1:1 allocation ratio. An independent statistician generated the randomization sequence using a computer-generated program with variable block sizes. Allocation concealment was ensured throughout the study. Participants, treating physicians, nurses, research personnel, and the statistician remained blinded to group assignment. After written informed consent was obtained, participants were assigned to their study group by the research nurse, who opened the next sequentially numbered opaque envelope. Each envelope contained two opaque syringes: one with ondansetron and one with placebo. The syringe labeled “1” was administered at triage. The second syringe was retained for potential rescue treatment and contained the alternate study medication (crossover design).

When a research nurse was available, children presenting to the ED with vomiting and/or diarrhea were identified by the triage nurse, who then notified a research assistant. The research assistant explained the study and assessed eligibility. After obtaining written informed consent, participants were randomized to one of the two study groups.

The treating physician determined whether the patient could be discharged immediately or required further observation for oral/IV rehydration or additional investigations.

Research assistants collected demographic data, reviewed medical charts for relevant variables, and obtained parent-completed comfort surveys. Follow-up telephone surveys were conducted 48– 72 hours after the intervention to evaluate secondary outcomes, including return visits and ongoing vomiting, including in children who left the ED without being seen. Children who left before physician assessment were excluded from the primary analysis but were contacted the following day to determine whether they sought care at another medical facility.

Data were entered into an Excel database (Microsoft Inc., Richmond, WA) and analyzed using SPSS version 25 (IBM Software Group Inc.). Baseline comparability between study arms was assessed using descriptive statistics: categorical variables were summarized as counts and percentages, while continuous variables were reported as mean, median, standard deviation, quartiles, and range (minimum–maximum).

The primary analysis compared the proportion of patients discharged immediately after the initial physician assessment between the ondansetron and placebo groups. A modified intention-to-treat approach was used, including all randomized patients who were seen by a physician. Length of stay and other continuous secondary outcomes such as the number of vomiting episodes in the ED and within 24 and 48 hours, and post-intervention comfort as measured by the BARF Scale (analyzed separately for children <7 years and ≥7 years) were analyzed using Student’s t-test. Binary secondary outcomes, including hospitalization, intravenous line insertion, incorrect diagnosis, and return visits to the ED within 48 hours or within one month, were analyzed using the Mantel-Haenszel test. An alpha level of 0.05 was applied for all analyses.

Prior to study initiation, we reviewed the medical records of 40 patients meeting the inclusion criteria. Based on this review, we estimated that 72.5% of eligible children would require observation after the first physician assessment in the control group. After consultation with multiple pediatric emergency physicians, we determined that a 20% absolute reduction in this proportion would be clinically meaningful. Assuming a power of 0.9 and α = 0.05, a total sample size of 248 patients (124 per arm) was required. Anticipating minimal loss to follow-up, we planned to recruit 260 children.

## Results

Between October 2018 and March 2020, 147 children were approached by a research nurse, of whom 124 met the inclusion/exclusion criteria and were invited to participate. Recruitment was slower than anticipated due to triage nurses being overwhelmed and unable to consistently notify the research team. The onset of the COVID-19 pandemic further disrupted ED operations, making triage-based research recruitment unfeasible. As a result, recruitment was prematurely stopped, and analysis was performed on the enrolled participants.

A total of 91 children (78%) consented and were randomized to receive either ondansetron (n = 44) or placebo (n = 47) immediately after triage. All participants were included in the primary analysis except one who left the ED before being seen by a physician (Figure 1). Baseline characteristics are summarized in Table 1. Most participants were aged 2–6 years, approximately half were male, and the majority presented with vomiting, with approximately 90% experiencing more than five episodes; diarrhea was uncommon at presentation. Median time from registration to physician assessment was 2.5 hours in both groups.

**Table 1.** Baseline characteristics of the study participants.

| Characteristics | Ondansetron<br>n=43 | Placebo<br>n= 47 |
| --- | --- | --- |
| Median age in months (1<sup>st</sup> and 3<sup>rd</sup> quartiles) | 36 (26, 75) | 53 (24, 72) |
| Median weight in Kg (1<sup>st</sup> and 3<sup>rd</sup> quartiles) | 15 (12, 21) | 15 (13, 23) |
| Sex male (%) | 24 (56) | 26 (55) |
| Vomiting in previous 24h<br>• 3-5<br>• 6-10<br>• >10 | • 6 (14)<br>• 20 (47)<br>• 17 (40) | • 3 (6)<br>• 19 (40)<br>• 25 (53) |
| Diarrhea in previous 24h<br>• 0<br>• 1-5<br>• 6-10<br>• >10 | • 31 (72)<br>• 8 (19)<br>• 3 (7)<br>• 1 (2) | • 32 (68)<br>• 11 (23)<br>• 2 (4)<br>• 1 (3) |
| Length of symptoms:<br>• 0-4h<br>• 4-<24h<br>• 24-<72h<br>• >= 72h | • 2 (7)<br>• 6 (19)<br>• 6 (19)<br>• 17 (55) | • 3 (10)<br>• 6 (19)<br>• 5 (16)<br>• 17 (55) |
| Median wait time in minutes to see physician (1<sup>st</sup> and 3<sup>rd</sup> quartiles) | 163 (125, 213) | 160 (126, 211) |
| Final diagnosis<br>• Gastro-enteritis<br>• Vomiting not specified<br>• Pharyngitis<br>• Other | • 31 (72)<br>• 6 (14)<br>• 1 (2)<br>• 5 (11) | • 37 (79)<br>• 7 (15)<br>• 1 (2)<br>• 2 (4) |

**Figure 1.**
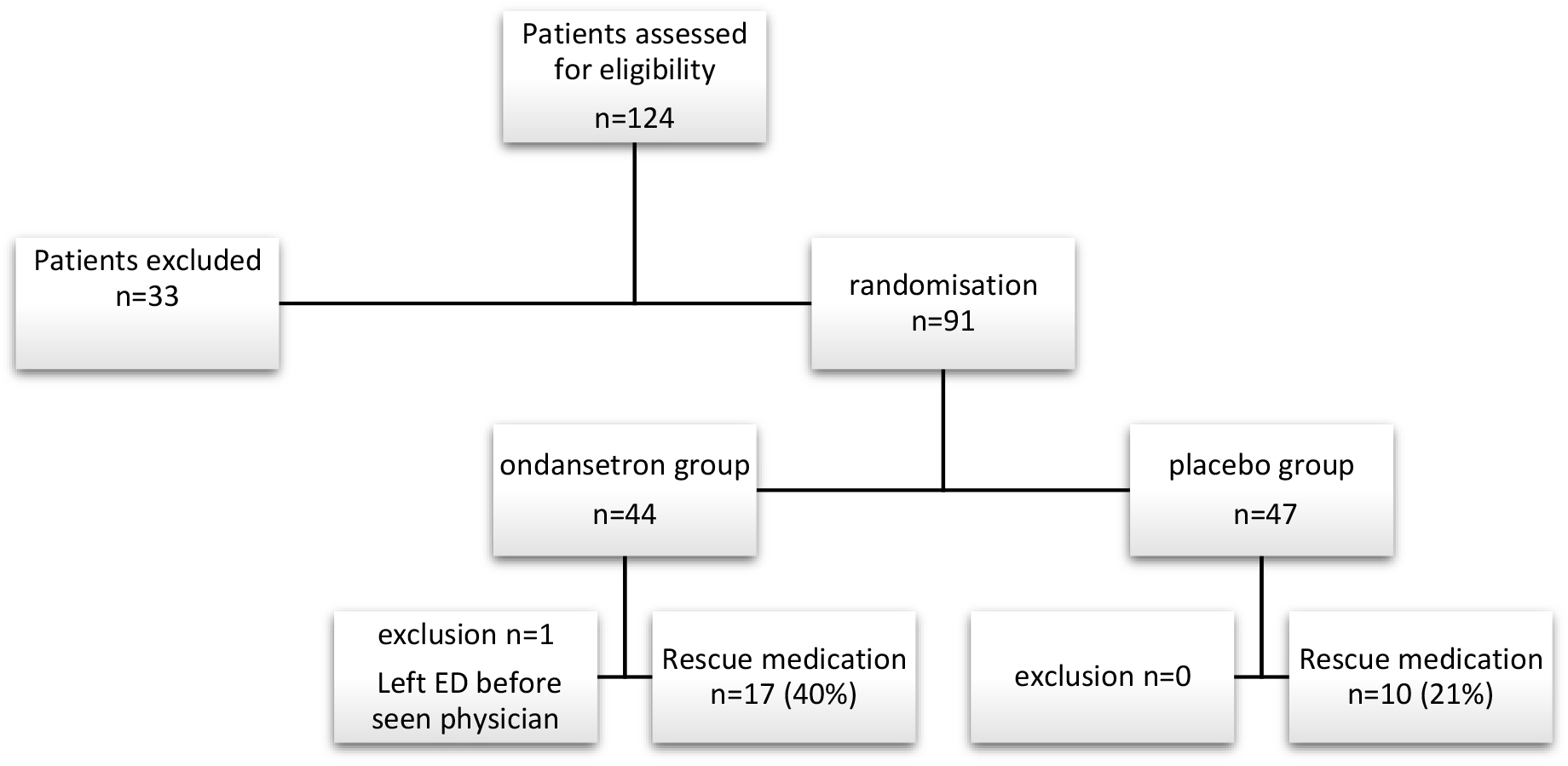
Flow diagram of the study participants

Forty participants (45%) were discharged immediately after the initial physician evaluation, with similar proportions in the ondansetron and placebo groups (19 [44%] vs. 21 [45%]; absolute difference 1%, 95% CI: −20% to 19%) (table 2). The mean volume of oral rehydration solution consumed between randomization and physician assessment was comparable (60 mL vs. 58 mL; difference 2 mL, 95% CI: −19 to 18 mL). No significant differences were observed in total ED length of stay (median 232 vs. 227 minutes, p = 0.677) or post-physician assessment length of stay (72 vs. 68 minutes, p = 0.821).

**Table 2.**
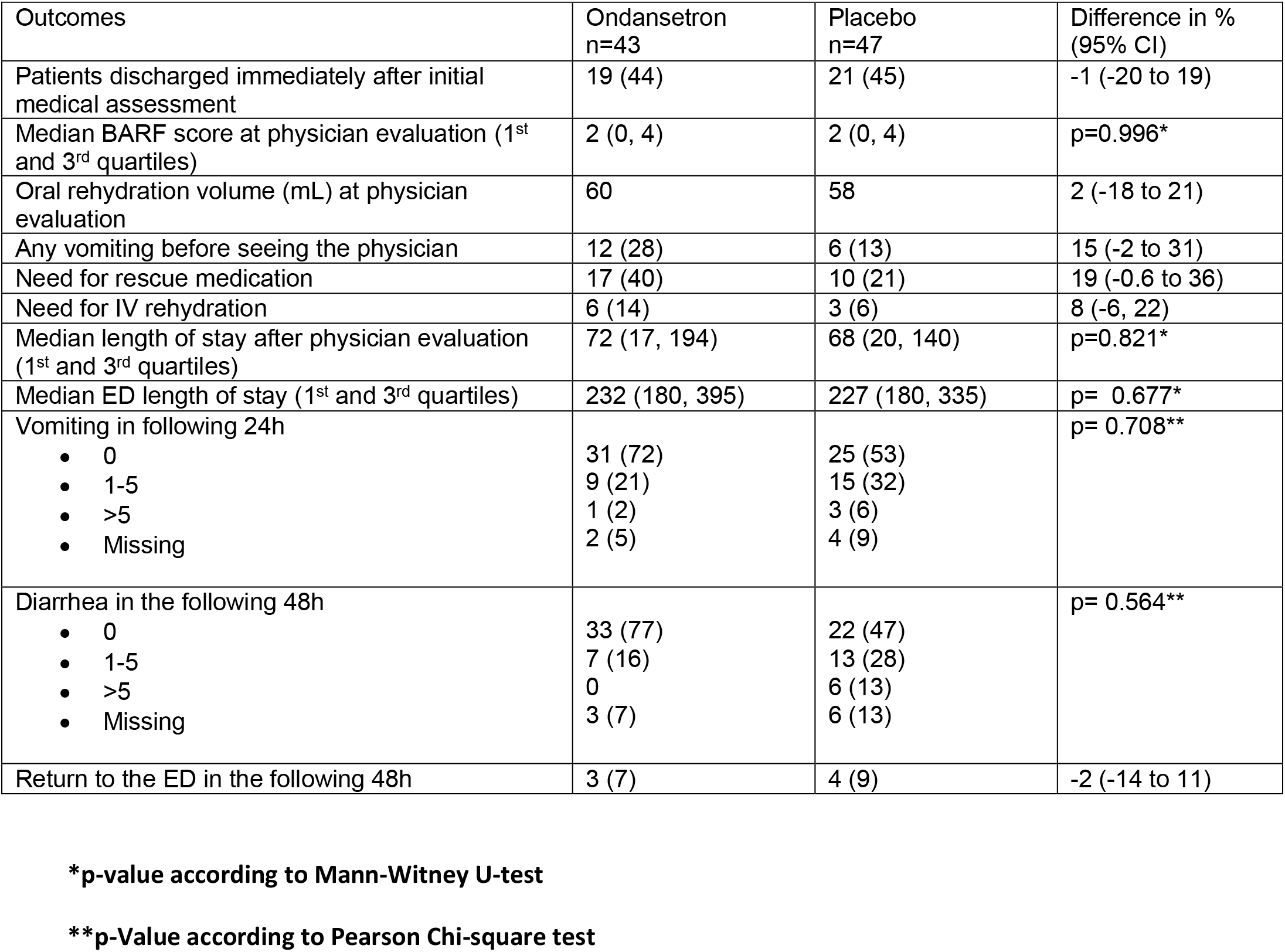
Results for the study participants.

A higher proportion of children in the ondansetron group reported vomiting after intervention while waiting for physician’s assessment (12 [28%] vs. 6 [13%]; difference 15%, 95% CI −2 to 31), but this did not reach statistical significance. There were no differences between groups in the need for rescue medication or intravenous rehydration (Table 2). There were also no significant differences in vomiting or diarrhea at 24–48 hours post-intervention. Return visits to the ED for persistent symptoms occurred in 3 (7%) and 4 (9%) participants in the ondansetron and placebo groups, respectively (difference 2%, 95% CI: −14 to 11).

A sub-analysis of the 63 children who waited more than 60 minutes between intervention and physician evaluation similarly found no differences in any outcomes (Web Appendix Table 2a).

Final diagnoses were gastroenteritis or unspecified vomiting in 37 (86%) and 44 (84%) children in the ondansetron and placebo groups (Table 1), respectively, confirming accurate triage nurse identification. No adverse effects were reported, and diarrhea within 48 hours did not differ between groups. Only one participant left the ED without being seen by a physician.

## Discussion

This prematurely stopped study failed to demonstrate a clinical benefit of initiating ondansetron at triage for children with AGE as children receiving ondansetron were not more likely to be discharged immediately than those receiving placebo. Also, there was no association between treatment groups and length of stay at the ED, amount of oral hydration drank by the participant, patient comfort or need of rescue medication.

To our knowledge, this is the first randomized trial evaluating the benefit of ondansetron at triage for children with AGE. A notable finding was the unexpectedly high discharge rate in the placebo group (45%). Previous internal data suggested that only 27% would be discharged immediately (73% requiring observation). Furthermore, the need for IV hydration was low in both groups (14% in ondansetron group vs 6% in placebo group). It was less than expected in the placebo group compared to the literature using similar inclusion/exclusion criteria. For example, Freedman and al. described that 14% of patients needed IV hydration in the ondansetron group and 31% in the placebo group^7^. In the Marchetti et al. study, IV hydration was needed in 11.8% of patients in the ondansetron group and 28.8% in the placebo group(10). In the Roslund et al. study, IV hydration was required in 21.6% of patients in the ondansetron group and 54.5% in the placebo group(8).

Several factors may explain our results. First, the presence of research assistants who coached families on oral rehydration therapy may have standardized and improved rehydration efforts. Outside the study, parents would have received standard instructions for oral rehydration at triage but without ongoing support until physician assessment. In the study, follow-up by the research assistant to assess the BARF scale, oral hydration, and vomiting may have inadvertently improved adherence to the hydration protocol, potentially enhancing outcomes in both groups. This could have improved outcome of participants in both groups, intervention and placebo. It is also possible that our patients had mild symptoms of AGE and did not really need medication to alleviate their symptoms. Finally, the use of a sucrose placebo might have had a metabolic effect, potentially alleviating ketosis-driven nausea(15, 16). Children with feeding intolerance often develop starvation ketosis and hypoglycemia, which can exacerbate nausea and vomiting. However, there is scarce medical literature supporting this hypothesis. A randomized double-blind study in 1976 compared glucose versus sucrose in addition to other electrolytes. The number of failures in the glucose-treated group was significantly greater than in the sucrose-treated group (27% vs. 10%; p=0.05)(15). Researchers in Egypt evaluated the effects of adding honey to oral hydration in a randomized controlled trial that involved 100 children with gastroenteritis. There was a significant reduction in vomiting (p < 0.001) and diarrhea frequency (p < 0.05) in the honey treated group compared with the control group (oral hydration alone)(17). Recent evidence suggests that starvation ketosis is highly prevalent in this population and that capillary test for ketones should be considered because these patients may require specific therapy(16). Sucrose placebo was chosen to best match the appearance and taste of ondansetron. Based on our results and these previous studies, evaluating the impact of oral sucrose prior to oral hydration in gastroenteritis could be of potential interest in a future study.

The study must be evaluated in the context of its limitations. First, the trial was significantly underpowered due to premature termination (91 of the planned 260 participants enrolled). Despite the smaller sample size, there was no trend suggesting a benefit of ondansetron, and the confidence interval did not exceed the 20% difference considered clinically meaningful by expert consensus. This suggests that the impact of triage-based administration may be less than anticipated. The decision to discharge a patient immediately after the initial physician assessment is inherently subjective and may depend on multiple factors. For example, a physician may choose to pursue additional investigations to confirm the diagnosis, even if the patient has stopped vomiting. Finally, the clinical relevance of the primary outcome, need for observation, is debatable. We chose this outcome instead of length of stay following advice from experts in pediatric emergency medicine who estimated that it would be less affected by external factors such as ED crowding.

In conclusion, this trial did not demonstrate a benefit of triage-initiated ondansetron in reducing the need for ED observation. However, the unexpectedly favorable outcomes in the sucrose placebo group underscore the importance of early, structured oral rehydration coaching. Future research should investigate whether targeted screening for starvation ketosis at triage and early administration of sucrose/glucose could provide a specific therapeutic pathway to resolve vomiting and enhance rehydration success in children with acute gastroenteritis.

## Data Availability

All data produced in the present study are available upon reasonable request to the authors

---

**Web appendix.** Table 2a Results for the study participants restricted to participants who received medication > 60 minutes before physician evaluation (n= 63)

| Outcomes | Ondansetron at triage<br>n=29 | Control<br>n=34 | Difference in %<br>(95% CI) |
| --- | --- | --- | --- |
| Patients discharged immediately after initial medical assessment | 16 (55) | 18 (53) | 2 (-21 to 25) |
| Median BARF score at physician evaluation (1<sup>st</sup> and 3<sup>rd</sup> quartiles) | 2 (0 and 4) | 2 (0 and 4) | p= 0.241* |
| Oral rehydration volume (in mL) at physician evaluation | 83 | 67 | 16 (-7 to 39) |
| Any vomiting before seeing the physician | 9 (31) | 4 (12) | 19 (-1 to 39) |
| Need for rescue medication | 13 (45) | 7 (21) | 19 (-- 1 to 39) |
| Need for IV rehydration | 5 (17) | 2 (6) | 11 (-5 to 29) |
| Median length of stay after physician evaluation (1<sup>st</sup> and 3<sup>rd</sup> quartiles) | 37 (14 and 119) | 55 (15 and 114) | p= 0.799* |
| Median ED length of stay (1<sup>st</sup> and 3<sup>rd</sup> quartiles) | 232 (195 and 395) | 237 (184 and 357) | p= 0.896* |
| Vomiting in following 24h<br>• 0<br>• 1-5<br>• >5<br>• Missing | 20 (69)<br>6 (21)<br>1 (3)<br>2 (7) | 18 (53)<br>10 (29)<br>3 (9)<br>3 (9) | p= 0.895** |
| Diarrhea in the following 48h<br>• 0<br>• 1-5<br>• >5<br>• Missing | 22 (76)<br>4 (14)<br>0<br>3 (10) | 15 (44)<br>10 (29)<br>5 (15)<br>4 (12) | p= 0.629** |
| Return to the ED in the following 48h | 1 (3) | 3 (9) | -5 (-20 to 10) |

\*p-value according to Mann-Witney U-test
\*\*p-Value according to Pearson Chi-square test

